# Associations of mitochondrial DNA copy number with incident risks of gastrointestinal cancers: a prospective case-cohort study

**DOI:** 10.1101/2021.12.25.21268390

**Authors:** Xin Guan, Mengying Li, Yansen Bai, Yue Feng, Guyanan Li, Wei Wei, Ming Fu, Hang Li, Chenming Wang, Jiali Jie, Hua Meng, Xiulong Wu, Qilin Deng, Fangqing Li, Handong Yang, Xiaomin Zhang, Meian He, Huan Guo

## Abstract

**Background:** Epidemiological investigations implied that inter-individual variations of mitochondrial DNA copy number (mtDNAcn) could trigger predisposition to multiple cancers, but evidence regarding gastrointestinal cancers (GICs) was still uncertain.

**Methods:** We conducted a case-cohort study within the prospective Dongfeng-Tongji cohort, including incident cases of colorectal cancer (CRC, n=278), gastric cancer (GC, n=138), and esophageal cancer (EC, n=72) as well as a random subcohort (n=1173), who were followed up from baseline to the end of 2018. Baseline blood mtDNAcn was determined with quantitative PCR assay, and associations of mtDNAcn with the GICs risks were estimated by using weighted Cox proportional hazards models.

**Results:** Significant *U*-shaped associations were observed between mtDNAcn and risks of CRC, GC, EC, and total GICs. Compared to subjects within the 2^nd^ quartile (Q2) mtDNAcn subgroup, those within the 1^st^ (Q1), 3^rd^ (Q3) and 4^th^ (Q4) quartile subgroups showed increased risks of CRC [HR(95%CI)=2.27(1.47-3.52), 1.65(1.04-2.62), and 2.81(1.85-4.28), respectively] and total GICs [HR(95%CI)=1.84(1.30-2.60), 1.47(1.03-2.10), and 2.51(1.82-3.47), respectively], and those within Q4 subgroup present elevated GC and EC risks [HR(95%CI)=2.16(1.31-3.54) and 2.38(1.13-5.02), respectively]. Similar associations of mtDNAcn with CRC and total GICs risks remained in stratified analyzes by age, gender, and smoking status. Notably, there were joint effects of age and smoking status with mtDNAcn on CRC and total GICs risks.

**Conclusions:** This prospectively case-cohort study showed *U*-shaped associations between mtDNAcn and incident risks of GICs, but further researches are needed to confirm these results and uncover underlying biological mechanisms.

## Introduction

Gastrointestinal cancers (GICs), including colorectal cancer (CRC), gastric cancer (GC), and esophageal cancer (EC), jointly account for over 22% of all cancer-related deaths and constitute over 21% and 13% of incident cancer cases among men and women, respectively (Sung et al. 2021). The incidences of GICs vary considerably by geographical regions, with approximately 40% of all cases occurring in China. Many risk factors contribute to the development of GICs, including chronic infection (e.g., *Helicobacter pylori*), lifestyle-related factors (e.g., alcohol drinking), and unhealthy diet, especially foods conserved with nitrates and nitrites (Brenner, Kloor, and Pox 2014, McCormack et al. 2017, Sung et al. 2021, Kumar et al. 2020, Smyth et al. 2020). However, because of the occult onset of GICs and a lack of specific screening methods for these cancers, GICs are always diagnosed at an advanced stage with poor survival outcomes (Brenner, Kloor, and Pox 2014, Codipilly et al. 2018, Zhang et al. 2018), which underlines the great demand to identify early biomarkers to discriminate high risk populations.

As a double-membrane organelle of eukaryotic cell, mitochondrion harbors its own DNA and plays a pivotal role in a series of biological process, including reactive oxygen species (ROS) and energy production (Lin and Beal 2006, Mishra and Chan 2016). Mitochondrial DNA copy number (mtDNAcn), a reflection of mitochondrial DNA (mtDNA) levels per cell, is a promising biomarker of mitochondrial function (Castellani et al. 2020). In fact, although the mtDNAcn varies among different cell types, the regulation of mtDNAcn is rigid and specific tissues/cells remain fairly stable mtDNAcn. Abnormal mtDNAcn variation induced by oxidative damage or inflammation was reported to be associated many chronic diseases, e.g. cardiovascular disease, neurodegenerative and metabolic disorders (Carew and Huang 2002, Lee and Wei 2005, Liu et al. 2003, Wu et al. 2017, Nunnari and Suomalainen 2012). More importantly, mtDNAcn alterations have also been observed in many types of cancers (e.g., breast cancer, prostate cancer), and may treated as a latent susceptible or diagnostic biomarker (Shen et al. 2015, Xu et al. 2020). Some epidemiological studies have investigated the associations of mtDNAcn with GICs but with inconsistent findings. Pilot data from two prior retrospective case-control studies in China and India revealed the positive association between high mtDNAcn and increased risk of CRC (Qu et al. 2011, Kumar et al. 2017), but another two CRC nested case-control studies in Shanghai Women’s Health Study (SWHS) and Nurse’s Health Study (NHS) reported inverse associations (Huang et al. 2014, Yang et al. 2019). Similar divergent results were also observed for the association of mtDNAcn with GC risk. An early case-control study (984 pairs) showed a positive relationship of mtDNAcn with GC risk (Zhu et al. 2017), but another nested case-control study of 162 GC cases and 299 controls reported null significant association (Liao et al. 2011). Only one small sample-sized (218 pairs) case-control study investigated the association of mtDNAcn with EC and found lower level of mtDNAcn in EC cases than in controls (Xu et al. 2013). Given the aforementioned conflicting findings, the relationships between mtDNAcn and GICs still need to be illustrated in larger prospective studies.

To the best of our knowledge, no prior studies have prospectively evaluated associations of mtDNAcn with GICs risks comprehensively. Here, we performed a prospective case-cohort study based on the Dongfeng-Tongji (DFTJ) cohort, including a random subcohort with 1173 subjects and 488 incident GICs (including 278 CRC, 138 GC, and 72 EC), to investigate the associations of blood mtDNAcn with incident risks of CRC, GC, EC, and total GICs.

## Materials and methods

### Study population

All subjects were participants from the DFTJ cohort, an ongoing population-based longitudinal study initiated in 2008 with enrollment of 27,009 retired employees from Dongfeng Motor Corporation (DMC) in Shiyan, Hubei, China. Details on the methodology for DFTJ cohort have been described previously (Wang et al. 2013). In brief, all participants voluntarily responded to the baseline questionnaires and completed medical examination during the period from September 2008 to June 2010. Meanwhile, a 15 mL of fasting venous blood sample donated by each subject was collected and sorted at -80°C before laboratory determinations.

The study protocol was ethically approved by the Ethics and Human Subject Committee of Tongji Medical College, Huazhong University of Science and Technology, and the written informed consent was obtained from each subject.

### Case-cohort study design and covariates assessment

Among the total 27,009 participants, after excluding those without available DNA samples (*n*=4784) or who had previous history of cancer (*n*=427) or who were lost to follow-up (*n*=140), the remained 21,658 participants were treated as the base cohort with the median follow-up time of 10.3 years. Subsequently, we randomly selected a representative subcohort of 1173 subjects within the base cohort by age- and gender-stratified sampling with an overall sampling rate of 5% (**Table S1**). Incident cases of GICs among the cohort population were identified by conducting the periodic record linkage analysis with the database from the local healthcare service systems, including five designated hospital of DMC, Social Insurance Centers, and the Center of Disease Control and Preventions. Cancer incidence data were available till Dec 31, 2018 and defined by professional physicians using the International Classification of Diseases, 10^th^ Revision (ICD-10). Among the 21,658 participants, we identified a total of 488 newly diagnosed GICs, including 278 incident CRC cases (C18.2-C20, 14 were in subcohort), 138 incident GC cases (C16.0-C16.9, 6 were in subcohort), and 72 incident EC cases (C15.0-C15.9, 5 were in subcohort). Hence, a total of 1636 participants were finally enrolled in this case-cohort study. The detailed selection criteria and cases distribution are shown in **Fig. 1**.

**Figure 1.**
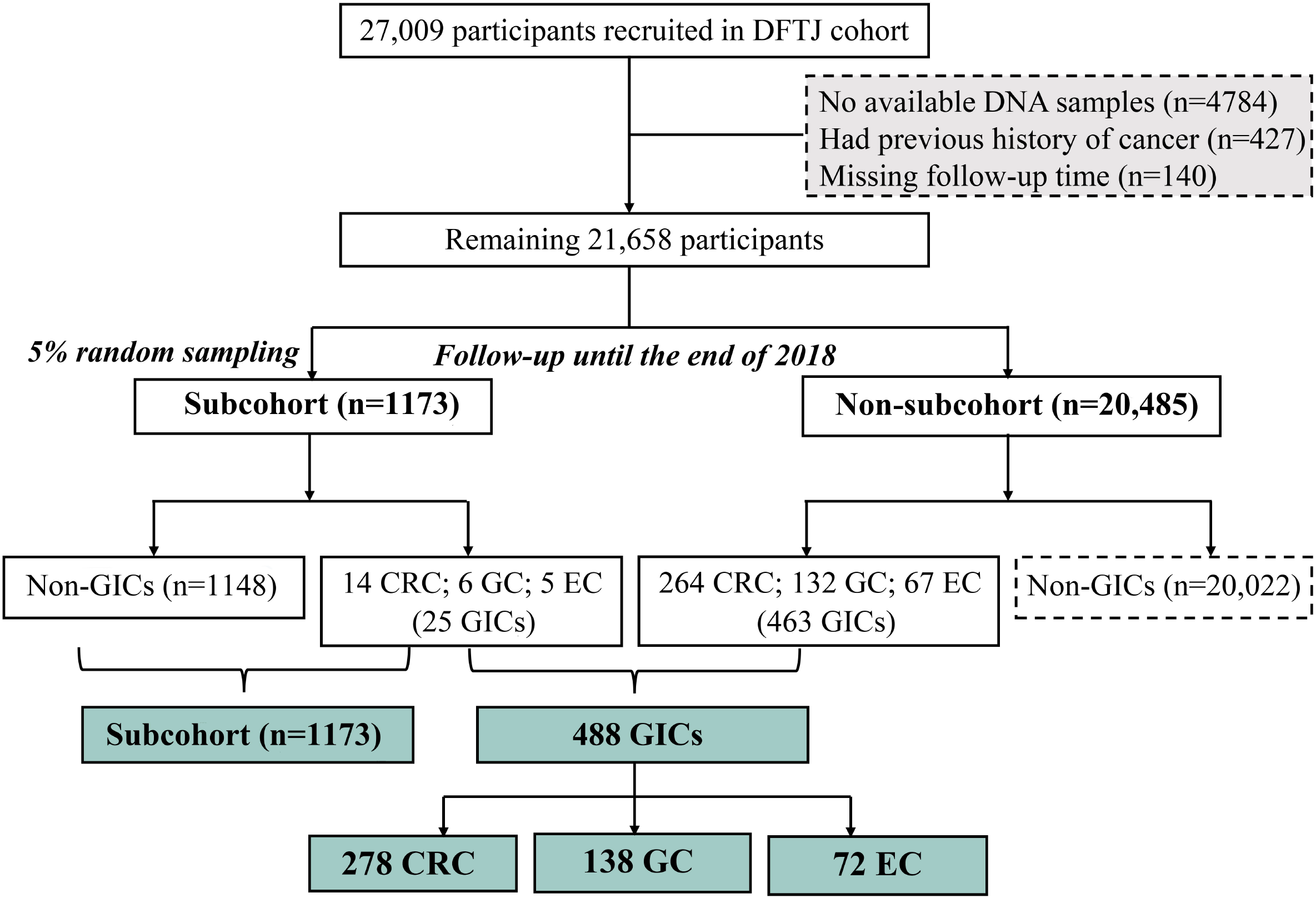
Case-cohort design nested within the DFTJ cohort. **Abbreviations:** DFTJ, Dongfeng-tongji; CRC, colorectal cancer; GC, gastric cancer; EC, esophageal cancer; GICs, gastrointestinal cancers.

The general information of all participants including demographic characteristics (e.g., age, gender, and education levels), lifestyles (status of smoking, alcohol drinking, and physical exercise), and chronic diseases history were recorded by conducting in-person interviews and completing structured questionnaires. Education achievements were coded as primary school or below, middle school, high school or beyond. Individuals who had smoked >1 cigarette per day for more than half a year were considered as current smokers and those who quitted smoking for more than six months were defined as former smokers, while the other subjects were never smokers. Participants who had drunk alcohol more than once a week for at least six months were defined as current alcohol drinkers, and former alcohol drinkers were those who had ever drunk alcohol but quitted for more than half a year. Otherwise, individuals were considered as never drinkers. Because of the low proportions of former smokers (<11.5%) and former drinkers (<6%) in the base cohort, we categorized current and former smokers into ever smokers, while former and never drinkers were merged into non-current drinkers. Regular physical exercises were considered as >20 min a time with at least five times per week and kept over half a year. The body mass index (BMI) was calculated according to the weight in kilograms divided by the square of height in meters (kg/m^2^). Basing on the first-degree familial information recorded, family history of cancer was defined as yes or no. The measurement of peripheral blood cells was conducted by using CELL-DYN 3700 (Abbott, USA) in accordance with standardized protocols.

### Determination of blood mtDNAcn at baseline

The total genomic DNA was extracted from peripheral blood samples by using Whole Blood DNA Extraction kit (BioTeke, Beijing, China). The quantitative real-time PCR (RT-PCR) was performed to measure relative mtDNAcn by using a high-throughput 384-well format with the QuantStudio™ 7 Flex Real-Time System (Applied Biosystems, CA, USA) as prior description with minor modifications (Janssen et al. 2012). In brief, each PCR reaction was conducted by wielding 1uL DNA sample (10 ng/uL) in a final volume of 10 uL per well. For mitochondrion amplification mixture, 1× PowerUp SYBR Green Master Mix (Applied Biosystems, 5uL/reaction), *ND1* primer [(300nmol/L FP 5′-ATGGCCAACCTCCTACTCCT-3′, 300nmol/L RP 5′-CTACAACGTTGGGGCCTTT -3′), (4 uL/reaction)] and RNase free water were contained, while the single-copy gene reaction mixture consisted of 1× PowerUp SYBR Green Master Mix (Applied Biosystems, 5 uL/reaction), *ACTB* primer [(300nmol/L FP 5′-ACTCTTCCAGCCTTCCTTCC-3′, 300nmol/L RP 5′-GGCAGGACTTAGCTTCCACA-3′), 4 uL/reaction] and RNase free water. The RT-PCR detection for *ND1* and *ACTB* copy number was assayed in triplicates in the same run, with thermal cycling profile proceeded at 50°C and 95 °C for 2 min respectively, followed by 40 cycles of 95°C for 15s and 60°C for 1min. The mtDNAcn was determined by using the ratio of mitochondrial gene (*ND1*) copy number to the single - copy reference gene (*ACTB*) copy number. For each 384-well plate, 200 DNA samples randomly selected from our study were equally pooled as the reference and subsequently serially diluted 1:2 to generate a six-point standard curve (SC) with concentration ranging from 1.25∼40 ng/μL, and R^2^ coefficient derived from the SC was >0.99. To ensure the quality of the measurement, the detection would be repeated if standard deviations (SD) of three cycle threshold (Ct) for any primers were greater than 0.30. Hence, the coefficient of variation (CV) calculated from the difference between *ND1* and *ACTB* Ct values presented preferable results that the inter- and intra-CV values were both less than 3%. To minimize the batch effects, all samples were tested randomly and values of mtDNAcn were normalized to the calibrator samples derived from the SC with concentration of 10 ng/uL.

### Statistical analysis

To explore the normality of continuous data, one-sample Kolmogorov-Smirnov test was performed. Mann-whitney *U* and Student’s *t*-test were conducted to estimate the difference of continuous variables between the incident GICs cases and controls in the subcohort, while Chi-square test was used for categorical data. Notably, since participants in the subcohort were free of cancer at baseline visit, for each specific subtype of GICs occurring, cases of the remaining cancer types in the subcohort were treated as controls (Pritchett et al. 2020). The mtDNAcn was log2-transformed to approach normality for further analysis. The associations of baseline characteristics with mtDNAcn were assessed by performing generalized linear models, with adjustment for age and gender.

Follow-up time was calculated from the date of enrollment to the date of GICs diagnosis, death, or December 31, 2018, whichever occurred first. Considering the difference of mtDNAcn between males and females, we categorized all participants into four subgroups according to the sex-specific quartiles of mtDNAcn in the subcohort subjects to evaluate the associations of mtDNAcn with incident risks of GICs (Thyagarajan et al. 2012). Subsequently, to handle with the oversampling of cases, the inverse probability weights and the robust sandwich variance estimators were performed, and adjusted hazard ratios (HRs) and 95% confidence intervals (CIs) were estimated by weighted Cox proportional hazards models (Barlow et al. 1999), with subjects in the 2^nd^ quartile (Q2) subgroup of mtDNAcn used as the reference group. Models were adjusted for age, gender, BMI, education levels (primary school or below, middle school, high school or beyond), smoking status (ever smokers/ non-smokers), alcohol drinking status (current/non-current drinkers), physical exercise (yes/no), and family history of cancer (yes/no). Furthermore, in order to explore the non-linear associations of mtDNAcn with GICs risks, the restricted cubic spline (RCS) regression models were conducted with knots at corresponding 5^th^, 50^th^, and 95^th^ percentiles, and the median of log2-transformed mtDNAcn was used as the reference value.

Next, we conducted stratified analysis by age (<65 y or ≥65 y), gender, and smoking status (ever smokers/ non-smokers) to explore the associations of mtDNAcn with CRC and total GICs risks in each stratum. The joint effects of mtDNAcn with age and smoking status were also evaluated. Given the comparative limited incident numbers of GC (*n*=138) and EC (*n*=72), the effects of mtDNAcn in aforementioned stratums were not evaluated for these two cancer types. To verify the robustness of our results, we subsequently implemented sensitivity analyses by adjusting for peripheral blood cells counts, or excluding participants diagnosed as GICs within the first two years of follow-up in the associations of mtDNAcn with GICs risks.

All statistical analyses were two-sided with *P* < 0.05 defined as statistically significant. SAS program (version 9.4, SAS Institute, Carry, NC) and R software (version 4.0.2) were used for data processing.

## Results

### General characteristics of study participants

Baseline characteristics, as well as levels of mtDNAcn, among the incident GICs cases and 1173 subcohort participants were presented in **Table 1**. The median follow-up period for the entire subcohort was 10.3 years. Compared to non-cases in the subcohort, the incident GICs cases were elder, more males and cigarette smokers, less educated (except for EC cases), and had a higher proportion of alcohol consumption (only for EC and total GICs cases) (All *P*<0.05). The incident cases of GC and total GICs both showed significantly higher level of mtDNAcn than the controls [median (interquartile range, IQR) of 1.00(0.78, 1.41) for GC and 1.00(0.73, 1.37) for total GICs v.s. 0.93(0.77, 1.15) for controls, both *P*<0.05)], while the levels of mtDNAcn in incident CRC and EC cases were suggestive elevated but did not reach the statistical significance (*P*=0.063 and 0.377, respectively).

**Table 1.** Baseline characteristics of subcohort participants and incident GICs cases.

| Characteristics <sup>a</sup> | Base cohort | Subcohort | Incident cases |  |  |  |
| --- | --- | --- | --- | --- | --- | --- |
|  |  |  | CRC | GC | EC | Total GICs <sup>b</sup> |
| Number of subjects | 21658 | 1173 | 278 | 138 | 72 | 488 |
| Age at enrollment, years | 63.2 ± 7.5 | 63.4 ± 7.7 | 67.2 ± 7.2** | 65.8 ± 7.5** | 65.7 ± 6.9* | 66.5 ± 7.3** |
| Males | 9653 (44.6) | 518 (44.2) | 162 (58.3)** | 83 (60.1)** | 55 (76.4)** | 300 (61.5)** |
| BMI, kg/m<sup>2</sup> | 24.5 ± 3.3 | 24.5 ± 3.2 | 24.9 ± 3.4 | 24.3 ± 3.3 | 23.7 ± 3.4* | 24.6 ± 3.4 |
| Education |  |  |  |  |  |  |
| Primary school or below | 6458 (29.8) | 331 (28.2) | 100 (36.0)* | 54 (39.1)* | 25 (34.7) | 179 (36.7)** |
| Middle school | 7943 (36.7) | 443 (37.8) | 97 (34.9) | 44 (31.9) | 28 (38.9) | 169 (34.6) |
| High school or beyond | 7257 (33.5) | 399 (34.0) | 81 (29.1) | 40 (29) | 19 (26.4) | 140 (28.7) |
| Smoking status |  |  |  |  |  |  |
| Never | 15296 (70.6) | 840 (71.6) | 174 (62.6)* | 78 (56.5)** | 27 (37.5)** | 279 (57.2)** |
| Former | 2469 (11.4) | 126 (10.7) | 39 (14.0) | 27 (19.6) | 16 (22.2) | 82 (16.8) |
| Current | 3893 (18.0) | 207 (17.7) | 65 (23.4) | 33 (23.9) | 29 (40.3) | 127 (26) |
| Alcohol drinking status |  |  |  |  |  |  |
| Never | 15796 (72.9) | 857 (73.0) | 201 (72.3) | 89 (64.5) | 35 (48.6)** | 325 (66.6)* |
| Former | 1167 (5.4) | 77 (6.6) | 22 (7.9) | 10 (7.2) | 5 (6.9) | 37 (7.6) |
| Current | 4695 (21.7) | 239 (20.4) | 55 (19.8) | 39 (28.3) | 32 (44.4) | 126 (25.8) |
| Physical activity, yes | 19286 (89.0) | 1061 (90.5) | 251 (90.3) | 117 (84.8)* | 62 (86.1) | 430 (88.1) |
| Family history of cancer, yes | 793 (3.7) | 37 (3.2) | 8 (2.9) | 2 (1.4) | 2 (2.8) | 12 (2.5) |
| Blood mtDNAcn | - | 0.93 (0.77, 1.15) | 1.01 (0.71, 1.35) | 1.00 (0.78, 1.41)* | 0.96 (0.77, 1.39) | 1.00 (0.73, 1.37)** |

**Abbreviations:** BMI, body mass index; mtDNAcn, mitochondrial DNA copy number; CRC, colorectal cancer; GC, gastric cancer; EC, esophageal cancer; GICs, gastrointestinal cancers.
**Notes:** Comparisons were made between incident cancer cases and non-case controls in the subcohort. In particular, when analyses were performed as for one specific type of cancer, cases of the remaining cancer types were treated as controls.
*P* values were derived from Student's *t*-test, Mann-Whitney *U* test, or Pearson's $\chi^2$ test. \*0.01<*P*<0.05; \*\**P*<0.01.
<sup>a</sup>Data were presented as mean ± SD, n (%), or median (25<sup>th</sup>, 75<sup>th</sup>). <sup>b</sup>Total GICs includes CRC, GC, and EC.

As **Fig. S1** depicted, the normality of mtDNAcn was improved by log2-transformation. Females had a significantly higher level of mtDNAcn than males [0.96(0.81, 1.18) *vs*. 0.89(0.72, 1.11), *P*<0.001]. The associations of baseline characteristics with mtDNAcn were presented in **Table S2**.

### Associations of mtDNAcn with GICs risks

The associations of blood mtDNAcn with GICs risks were summarized in **Table 2**. Compared to subjects within the Q2 subgroup of mtDNAcn, there was a significantly elevated risk of incident CRC for those within the 1^st^ (Q1), 3^rd^ (Q3) and 4^th^ (Q4) quartile subgroups of mtDNAcn [HR(95%CI)=2.27(1.47, 3.52), 1.65(1.04, 2.62) and 2.81(1.85,4.28), respectively]. Similar results were also observed for the associations of Q1, Q3 and Q4 mtDNAcn subgroups with incident risk of total GICs when compared to the Q2 mtDNAcn subgroup, with HR(95%CI) presented as 1.84(1.30, 2.60), 1.47(1.03, 2.10), and 2.51(1.82, 3.47) respectively. However, only the Q4 subgroup of mtDNAcn revealed significantly increased risks of GC and EC with the comparison of Q2 subgroup [HR(95%CI)=2.16 (1.31, 3.54) and 2.38(1.13, 5.02), respectively]. We further estimated the non-linear correlations between mtDNAcn and GICs risks by using RCS function, and observed the evident *U*-shaped associations of mtDNAcn with incident risks of CRC, GC, EC, and total GICs (all *P* for non-linear associations <0.05, **Fig. 2**). The above findings were not materially altered after further adjustment for platelet and white blood cell counts (**Table S3**), or excluding the incident GICs cases diagnosed within two years of follow-up, or excluding the other cancer cases when performing analysis for one specific type of GICs (**Table 2**).

**Table 2.** Associations of blood mtDNAcn with incident risks of GICs.

| Incident GICs | Quartiles of blood mtDNAcn <sup>a</sup> |  |  |  |
| --- | --- | --- | --- | --- |
|  | Q1 | Q2 | Q3 | Q4 |
| **CRC** |  |  |  |  |
| *All subjects* |  |  |  |  |
| No. of cases / subcohort | 83 / 295 | 37 / 292 | 56 / 292 | 102 / 294 |
| HR (95% CI)<sup>b</sup> | 2.27 (1.47, 3.52) | reference | 1.65 (1.04, 2.62) | 2.81 (1.85, 4.28) |
| *Excluding subjects who diagnosed during first two years of follow-up* |  |  |  |  |
| No. of cases / subcohort | 73 / 295 | 37 / 292 | 50 / 292 | 89 / 294 |
| HR (95% CI)<sup>b</sup> | 2.00 (1.28, 3.12) | reference | 1.47 (0.92, 2.35) | 2.47 (1.61, 3.78) |
| *Excluding other incident cancer cases* |  |  |  |  |
| No. of cases / subcohort | 83 / 272 | 37 / 280 | 56 / 277 | 102 / 263 |
| HR (95% CI)<sup>b</sup> | 2.31 (1.49, 3.59) | reference | 1.69 (1.07, 2.69) | 2.95 (1.93, 4.51) |
| **GC** |  |  |  |  |
| *All subjects* |  |  |  |  |
| No. of cases / subcohort | 32 / 295 | 26 / 292 | 26 / 292 | 54 / 294 |
| HR (95% CI)<sup>b</sup> | 1.28 (0.73, 2.24) | reference | 1.10 (0.61, 1.97) | 2.16 (1.31, 3.54) |
| *Excluding subjects who diagnosed during first two years of follow-up* |  |  |  |  |
| No. of cases / subcohort | 29 / 295 | 23 / 292 | 23 / 292 | 46 / 294 |
| HR (95% CI)<sup>b</sup> | 1.33 (0.73, 2.40) | reference | 1.11 (0.60, 2.06) | 2.10 (1.24, 3.56) |
| *Excluding other incident cancer cases* |  |  |  |  |
| No. of cases / subcohort | 32 / 267 | 26 / 281 | 26 / 274 | 54 / 262 |
| HR (95% CI)<sup>b</sup> | 1.32 (0.75, 2.32) | reference | 1.15 (0.64, 2.06) | 2.30 (1.40, 3.80) |
| **EC** |  |  |  |  |
| *All subjects* |  |  |  |  |
| No. of cases / subcohort | 18 / 295 | 11 / 292 | 18 / 292 | 25 / 294 |
| HR (95% CI)<sup>b</sup> | 1.75 (0.80, 3.85) | reference | 1.82 (0.82, 4.04) | 2.38 (1.13, 5.02) |
| *Excluding subjects who diagnosed during first two years of follow-up* |  |  |  |  |
| No. of cases / subcohort | 14 / 295 | 11 / 292 | 14 / 292 | 22 / 294 |
| HR (95% CI)<sup>b</sup> | 1.36 (0.59, 3.13) | reference | 1.46 (0.62, 3.42) | 2.13 (0.99, 4.57) |
| *Excluding other incident cancer cases* |  |  |  |  |
| No. of cases / subcohort | 18 / 268 | 11 / 278 | 18 / 275 | 25 / 262 |
| HR (95% CI)<sup>b</sup> | 1.75 (0.80, 3.86) | reference | 1.88 (0.84, 4.22) | 2.57 (1.22, 5.45) |
| **Total GICs** |  |  |  |  |
| *All subjects* |  |  |  |  |
| No. of cases / subcohort | 133 / 295 | 74 / 292 | 100 / 292 | 181 / 294 |
| HR (95% CI)<sup>b</sup> | 1.84 (1.30, 2.60) | reference | 1.47 (1.03, 2.10) | 2.51 (1.82, 3.47) |
| *Excluding subjects who diagnosed during first two years of follow-up* |  |  |  |  |
| No. of cases / subcohort | 116 / 295 | 71 / 292 | 87 / 292 | 157 / 294 |
| HR (95% CI)<sup>b</sup> | 1.68 (1.18, 2.40) | reference | 1.34 (0.93, 1.94) | 2.88 (1.64, 3.19) |
| *Excluding other incident cancer cases* |  |  |  |  |
| No. of cases / subcohort | 133 / 273 | 74 / 283 | 100 / 278 | 81 / 269 |
| HR (95% CI)<sup>b</sup> | 1.89 (1.34, 2.68) | reference | 1.52 (1.06, 2.17) | 2.64 (1.90, 3.66) |

**Abbreviations:** HR, hazard ratio; CI, confidence interval; mtDNAcn, mitochondrial DNA copy number; CRC, colorectal cancer; GC, gastric cancer; EC, esophageal cancer; GICs, gastrointestinal cancers.
**Notes:** <sup>a</sup> Quartile groups were divided by sex-specific cut-off values of mtDNAcn among sub-cohort participants. For males: <0.722 (Q1), 0.722-0.887 (Q2), 0.888-1.110 (Q3), $\geq 1.11$ (Q4); for females: <0.806 (Q1), 0.806-0.957 (Q2), 0.958-1.179 (Q3), $\geq 1.180$ (Q4).
<sup>b</sup> Weighted Cox proportional hazards models, with adjustment for age, BMI, gender, smoking (smokers/never smokers), alcohol drinking (current/non-current alcohol drinkers), physical activity (yes/no), education status (primary school or below, middle school, high school or beyond), and family history of cancer.

**Figure 2.**
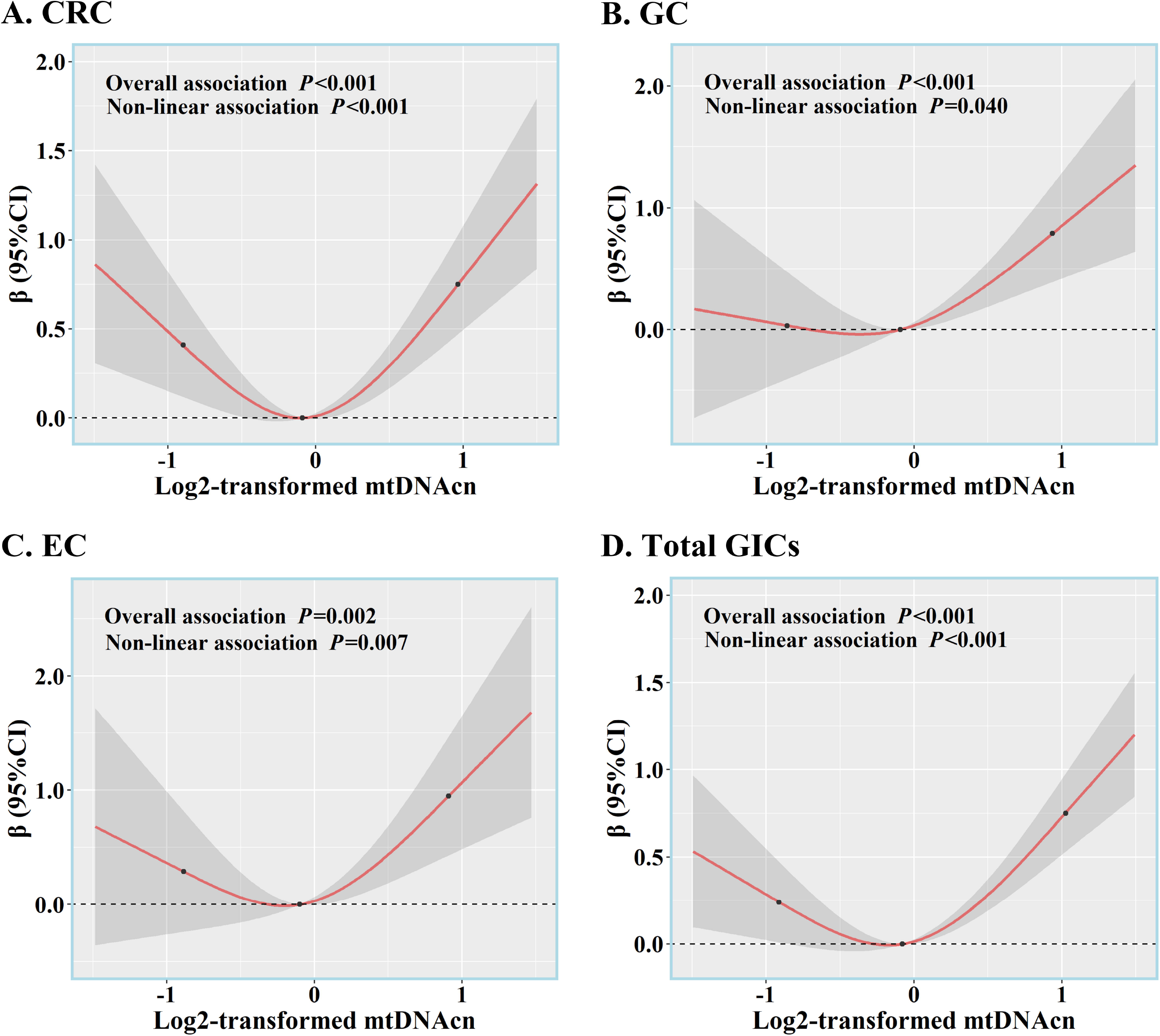
Associations of blood mtDNAcn with incident risks of GICs based on the restricted cubic spline function. (A) Association between mtDNAcn and incident risk of CRC. (B) Association between mtDNAcn and incident risk of GC. (C) Association between mtDNAcn and incident risk of EC. (D) Association between mtDNAcn and incident risk of total GICs. **Abbreviations:** mtDNAcn, mitochondrial DNA copy number; CRC, colorectal cancer; GC, gastric cancer; EC, esophageal cancer; GICs, gastrointestinal cancers. **Notes:** The three black solid dots represent the knots of 5^th^, 50^th^ and 95^th^ percentiles of log2, transformed mtDNAcn, while the separate median value of log2-transformed mtDNAcn was used as the reference.

### Stratification analysis

Since there were limited numbers of GC (n=138) and EC (n=72) cases, the further stratification analyses by age, gender, and smoking status were only conducted for CRC and total GICs. Individuals with age <65 years old showed increased risks of CRC and total GICs in Q1 and Q4 mtDNAcn subgroups when compared with the Q2 subgroup [CRC: HR(95%CI) = 2.34(1.23, 4.46) and 2.58(1.36, 4.88); total GICs: HR(95%CI)=1.95(1.20, 3.18) and 2.22(1.38, 3.56)] (**Fig. 3A**). We also observed elevated risks of CRC [v.s. Q2 subgroup of mtDNAcn, HR(95%CI)=2.19(1.22, 3.92) for Q1 and 2.84(1.63, 4.94) for Q4 subgroups] and total GICs [v.s. Q2 subgroup of mtDNAcn, HR(95%CI)=1.80(1.11, 2.91), 1.82(1.10, 3.04)], and 2.71(1.73, 4.25) for Q1, Q3, and Q4 subgroups] among participants with age ≥65 (**Fig. 3A**).

**Figure 3.**
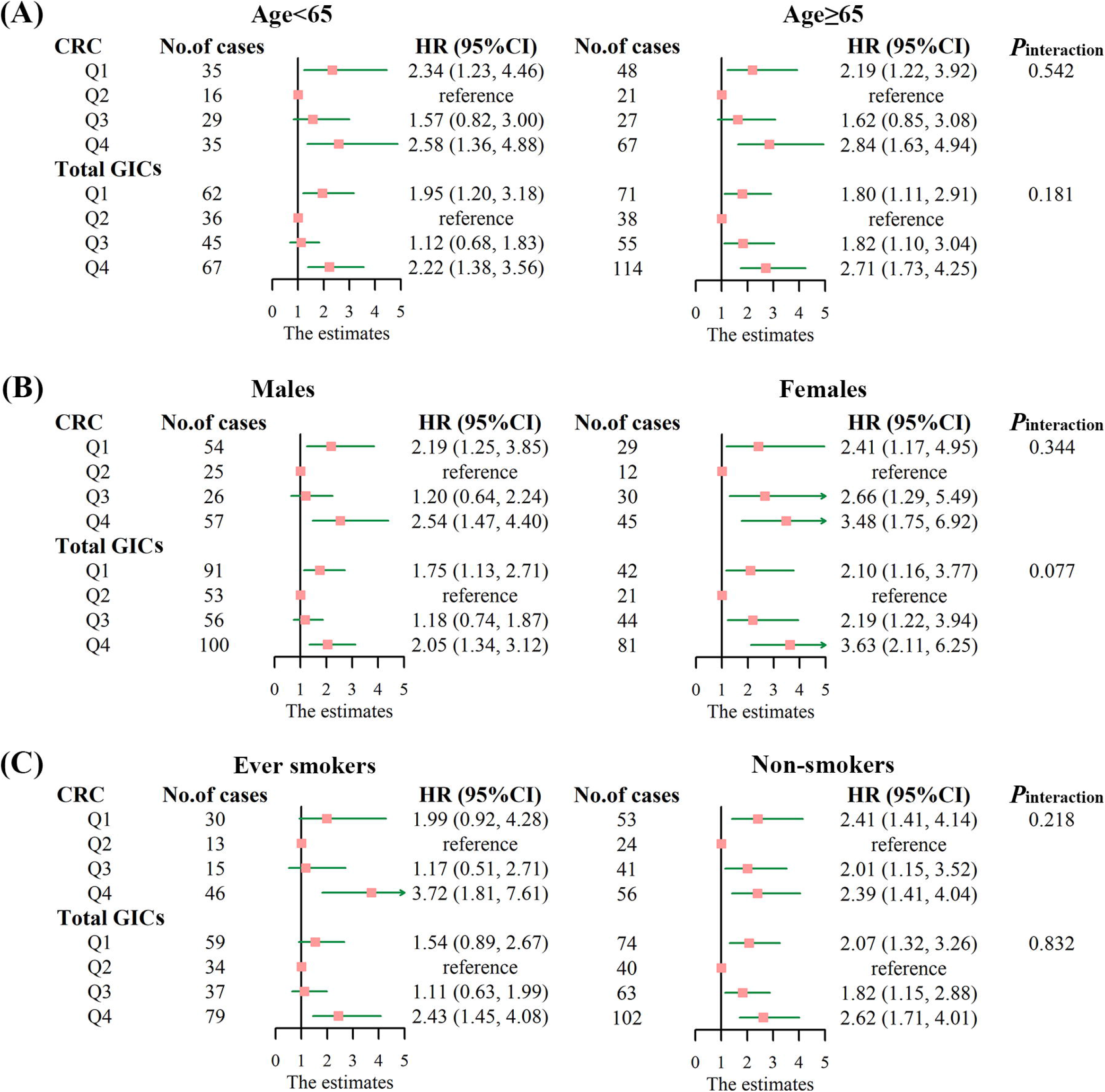
Stratified analyses for the associations of blood mtDNAcn with CRC and total GICs risk by age, gender, and smoking status. (A) Stratified by age; (B) Stratified by gender; (C) Stratified by smoking status. **Abbreviations:** HR, hazard ratio; CI, confidence interval; mtDNAcn, mitochondrial DNA copy number; CRC, colorectal cancer; GC, gastric cancer; EC, esophageal cancer; GICs, gastrointestinal cancers. **Notes:** Weighted Cox proportional hazards models, with adjustments for age, BMI, gender, smoking status, alcohol drinking status, physical exercise, education status, and family history of cancer, except for the stratification variable. The pink solid square and green whisker line in panels represent hazard ratios (HRs) and 95% confidence intervals (CIs), respectively. The green arrow means that the 95%CI exceeds the limitation of axis and not shown in this figure.

Among males, with the comparison of Q2 mtDNAcn subgroup, the Q1 and Q4 subgroups consistently showed raised risks of CRC [Q1: HR(95%CI)=2.19(1.25, 3.85), Q4: HR(95%CI)=2.54(1.47, 4.40)] and total GICs [Q1: HR(95%CI)=1.75(1.13, 2.71), Q4: HR(95%CI)=2.05(1.34, 3.12)], while the corresponding effects were strengthened among females that the Q1, Q3 and Q4 mtDNAcn subgroups presented 2.41-fold (95%CI: 1.17, 4.95), 2.66-fold (95%CI: 1.29, 5.49), and 3.48-fold (95%CI: 1.75, 6.92) risk of CRC, and 2.10-fold (95%CI: 1.16, 3.77), 2.19-fold (95%CI: 1.22, 3.94), and 3.63-fold (95%CI: 2.11, 6.25) risk of total GICs (**Fig. 3B**).

Among ever smokers, we only observed elevated risks of CRC and total GICs in Q4 mtDNAcn subgroups when compared with the Q2 subgroup [HR (95% CI)=3.72(1.81, 7.61) and 2.43(1.45, 4.08), respectively] (**Fig. 3C**). Among non-smokers, the significant increased risks of CRC and total GICs were observed among Q1, Q3, and Q4 subgroups [CRC: HR(95%CI)=2.41(1.41, 4.14), 2.01(1.15, 3.52), 2.39(1.41, 4.04), respectively; GICs: HR(95%CI)=2.07(1.32, 3.26), 1.82(1.15, 2.88), 2.62(1.17, 4.01), respectively] (**Fig. 3C**). However, we did not observe the modification effects of age, gender, and smoking status on the associations of mtDNAcn with CRC and total GICs risks (all *P*_interaction_ > 0.05, **Fig. 3**).

### Joint effects of mtDNAcn with age and smoking status

We further investigate the joint effects of mtDNAcn with age and smoking status on the incident risks of CRC and total GICs. When using subjects aged < 65 and within the Q2 mtDNAcn subgroup as the reference group, there were significantly increased incident risk of CRC [HR(95%CI)=3.75(2.01, 6.99), 2.71(1.37, 5.37), and 4.89(2.69, 8.92)] and total GICs [HR(95%CI)=2.51(1.56, 4.03), 2.52(1.52, 4.17), and 3.78(2.43, 5.90), respectively] among subjects with age ≥65 and within Q1, Q3, and Q4 mtDNAcn subgroups (**Fig. 4**). Compared to non-smokers within Q2 mtDNAcn subgroup, we also observed significant increased risk of CRC among ever smokers within Q1 and Q4 mtDNAcn subgroups [HR(95%CI)=2.54(1.32, 4.91) and 4.48(2.39, 8.37), respectively], and increased risk of total GICs among ever smokers within Q1, Q3, and Q4 subgroups [HR(95%CI)=2.67(1.58, 4.53), 1.88(1.08, 3.28), and 4.10(2.46, 6.81), respectively] (**Fig. 4**).

**Figure 4.**
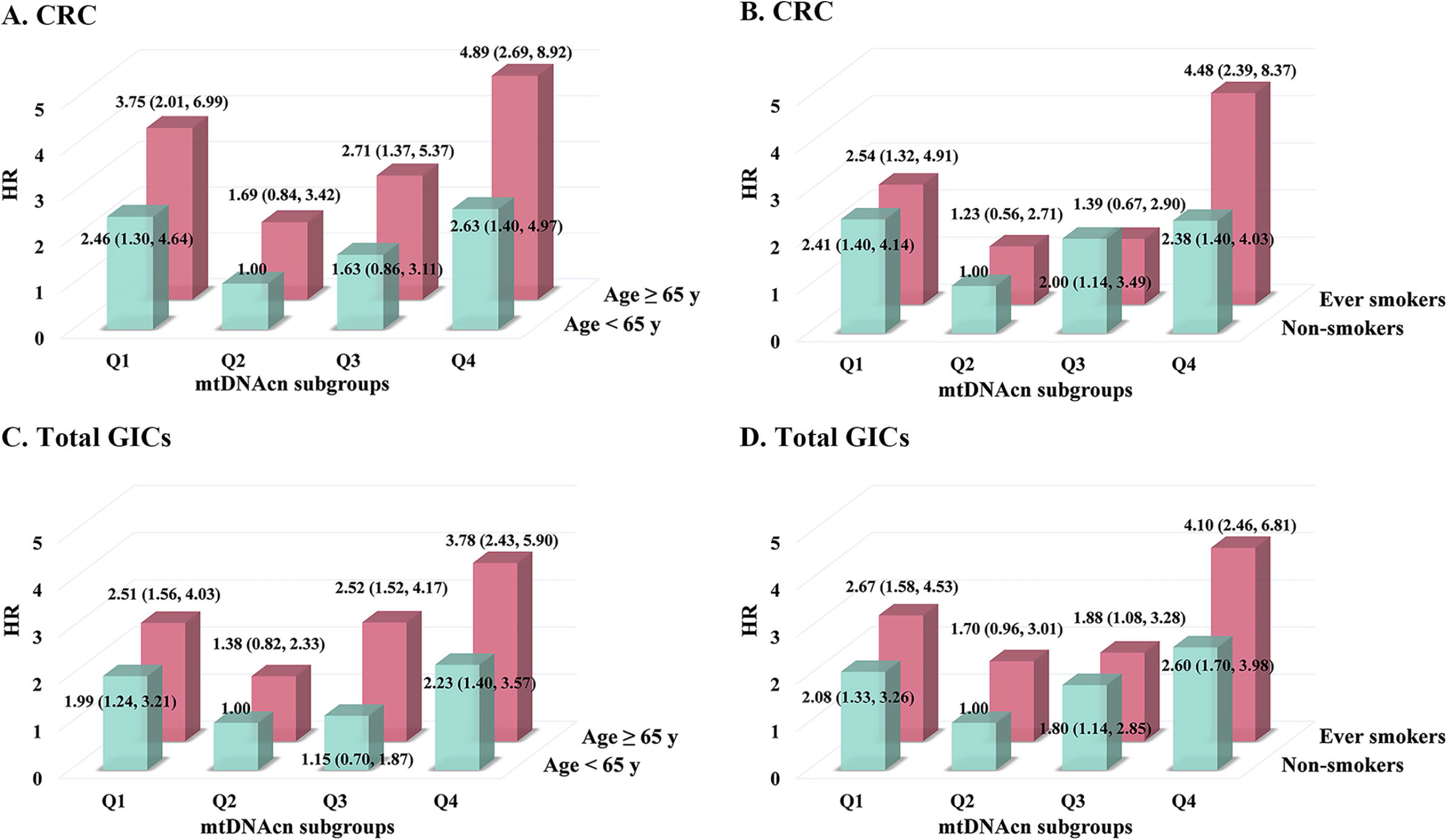
Joint effects of blood mtDNAcn with age and smoking status on CRC and total GICs risks. (A) Joint effect of mtDNAcn with age on CRC. (B) Joint effect of mtDNAcn with smoking status on CRC. (C) Joint effect of mtDNAcn with age on total GICs. (D) Joint effect of mtDNAcn with smoking status on total GICs. **Abbreviations:** HR, hazard ratio; CI, confidence intervals; mtDNAcn, mitochondrial DNA copy number; CRC, colorectal cancer; GICs, gastrointestinal cancers. **Notes:** Weighted Cox proportional hazards models, with adjustments for age, BMI, gender, smoking status, alcohol drinking status, physical exercise, education status, and family history of cancer, except for variables in the combined terms.

## Discussion

In this prospective case-cohort study, we observed that comparatively lower and higher levels of blood mtDNAcn were associated with 47% to 181% increased incident risks of GICs. The nonlinear *U*-shaped associations of mtDNAcn with CRC, GC, EC and total GICs were further manifested by the RCS curves. In addition, we observed the joint effects of mtDNAcn with elder age (≥65 years-old) and tobacco smoking on increased risks of CRC and total GICs. These findings provided novel prospective epidemiological evidence for the effect of mtDNAcn on the development of GICs.

Chronic accumulation in oxygen radical generation in mitochondria could result in catastrophic cycle of mtDNA damage as well as functional impairment. This would promote ROS (e.g., superoxide and hydroxyl radicals) production, and cause cellular defective apoptosis regulation, which might generate increased nuclear DNA damage and potentiated carcinogenesis (Chan 2020, Idelchik et al. 2017). Recently, increasing studies have assessed that either low or high level of mtDNAcn was correlated with elevated risk of cancer in type-specific manner (Sun et al. 2016, Zheng et al. 2019, He et al. 2014). However, relevant epidemiological evidence on the associations between mtDNAcn and GICs is limited. In this prospective case-cohort study, we observed significant nonlinear *U*-shaped associations of blood mtDNAcn with the developments of CRC, GC, and EC, which indicated that individuals with low and high mtDNAcn had elevated incident risks of GICs. To be consistent with our results, Thyagarajan et al. also observed a significant *U*-shaped relationship between mtDNAcn and CRC risk (*P*_curvilinearity_ < 0.001) in a nested case-control study of 422 CRC cases and 874 controls, while this effect was presented in both men and women participants (Thyagarajan et al. 2012). But another Chinese nested case-control study did not observe the significant association between mtDNAcn and GC risk (Liao et al. 2011, Sun et al. 2014). Relevant population-based evidence for the association between mtDNAcn and EC risk was actually scanty. Hence, our finding of *U*-shaped pattern in the relationship between mtDNAcn and EC risk is novel and could be an objective supplement of prospective epidemiological evidence. In general, the contradictory in reported relationship directions between mtDNA and GICs risks could be partially attributed to the nonlinear *U*-shaped pattern, as well as the population select bias, sample size limitation, random chance, and reverse causality. Therefore, larger and well-designed studies, especially the longitudinal cohort designs, are warranted to replicate our findings in different ethnic populations.

The mechanisms underlying the associations of blood mtDNAcn with GICs etiology are not completely understood, but some existed evidence demonstrates that the *U*-shaped associations are biologically plausible. Mitochondria play a vital role in the evolution of tumor cell genome, acting as a double-edged sword in cancer developing. An *in vitro* study found that the H_2_O_2_ treated lung fibroblast (MRC-5) could be blocked at G(0) and G(1) phases accompanying with ROS generation, and further mtDNA content was significantly elevated in the concentration and time dependent manner, indicating that mild oxidative stress may trigger increase of mtDNA through the cell-cycle arrest pathway (Lee et al. 2000). Also, one previous two-stage case-control study reported that high level of 8-hydroxy-2’-deoxyguanosine (8-OHdG) (Zheng et al. 2019), an widely used biomarker of DNA oxidative damage and well documented in GICs development(Guo et al. 2016, Ma et al. 2013), were associated with increased mtDNAcn. Another two independent studies revealed that mtDNAcn was significantly higher in tumor tissue of CRC patients than the corresponding normal tissues (Lim et al. 2012, Gao et al. 2015). These findings propose the hypothesis that ROS might induce mtDNAcn and further cause more oxidative damage to intracellular DNA and other constituents, which might accelerate the initiation and progression of tumorigenesis (Dizdaroglu 2015). Whereas, it should not be neglect that increased mtDNAcn generated from ageing cells as a result of the feedback response compensating for aberrant mitochondria respiratory chain or mtDNAcn mutation (Lee and Wei 2005), might mask the association between elevated mtDNAcn and raised risks of GICs.

Once the rate of oxidative injury overwhelms the capacity of the compensation mechanism of mtDNA, the impermanent increased mtDNAcn can no longer deal with the oxidative stress, and followed by eventual attrition of mtDNA that would trigger a net decrease in mtDNAcn, since the mtDNA was degraded by the intracellular enzyme system to prevent excessive accumulation damage of oxidative stress (Shokolenko et al. 2009, Lee and Wei 2005). A prior *in vitro* study observed that a decrease in mitochondria in HeLa cells would result in raised lipid peroxidation and reduced activity of antioxidant enzyme (e.g., catalase), and further induced elevated DNA damage in response to the oxidative stress (Delsite et al. 2003). More recently, an *in vivo* study conducted among mice revealed that impaired mitochondrial would release oxidized mtDNA into cytosol and subsequently activate the inflammasomes (Shimada et al. 2012), which can trigger the production of proinflammatory cytokines including interleukin-1β (IL-1β) and interleukin-18 (IL-18) (Zhong, Liang, and Zhong 2019). Consistent with the vital role of the inflammasomes in activating inflammatory reaction, high levels of released IL-1β and IL-18 have been correlated with increased risks of CRC and GC (Kolb et al. 2014, Haghshenas et al. 2009). In general, the aforementioned findings support the plausible biologic mechanisms by which the lower mtDNAcn might be related with elevated risks of GICs, but the pathogenesis effects of mtDNAcn still need validations and further in-depth investigations.

Since the gastrointestinal system owned similar mucosa epithelial barrier, the homeostasis of epithelial cell played a vital role in the normal gastrointestinal functional progression (Shalapour and Karin 2020). It should be realized that mitochondrial dysfunction and mtDNA variation may cause abnormal epithelium energy metabolism, which would lead to the loss of epithelial barrier integrity (Hu et al. 2018). Once the protective barrier was compromised, numerous microbes and their products would result in activation of inflammation factor NF-κB and induction of IL-6 and TNF, which can promote the development of colon and liver cancers (Greten et al. 2004, Pikarsky et al. 2004). These roles might explain the consistent effects of mtDNAcn on CRC, GC, and EC incident risks found in our study. Also, the present study showed robust *U*-shaped association of mtDNAcn with CRC and total GICs risks among subjects with the strata of age, smoking status, and gender, which were consistent with two prior studies (Thyagarajan et al. 2012, Wang et al. 2018). Especially, our findings of more robust effects among women than men were in line with previous results (Thyagarajan et al. 2012). One possible mechanism might because females seem be more susceptible to oxidative damage induced by hormone stimulation. As shown in experimentally human breast cancer cell lines MCF7, estrogen exposure revealed increased mitochondrial ROS production, suggesting that estrogen might contribute to susceptibility of oxidative injury among females (Parkash, Felty, and Roy 2006). Additionally, we observed the combined effect of mtDNAcn with age and smoking status on risks of developing CRC and total GICs, which might result from the shared features of increasing ROS by aging and smoking, as well as mitochondrial dysfunction (Parajuli et al. 2013, Liguori et al. 2018, Shokolenko et al. 2009). Nevertheless, the associations of blood mtDNAcn with incident risks of CRC and total GICs could not be modified by age, gender, and smoking status, indicating that mtDNAcn was an independent risk factor.

This study was the first to provide prospective epidemiological evidence on the nonlinear *U*-shaped associations of mtDNAcn with GC, EC, and total GICs risks, as well as validating *U*-shaped pattern between mtDNAcn and CRC risk reported in a prior nested-case control study (Thyagarajan et al. 2012). The prospective design here owns more power to illuminate time order and causal relationships than the case-control designs. Additionally, the case-cohort analysis including three types of GICs can be performed with the same comparison group, improving the statistical efficiency and coping with multiple outcomes of interest (Barlow et al. 1999). Since large longitudinal designs with few observed outcomes might require massive resources to identify covariate information, the case-cohort frame can complete data collection with less cost and be more flexibility, providing both a window on the entire cohort and the chance to evaluate quality control (Barlow et al. 1999). Meanwhile, after excluding GICs diagnosed within 2 years of cohort entry or adjusting possible confounds (e.g., platelet and white blood cell counts), the *U*-shaped pattern still existed, indicating the robust associations between mtDNAcn and GICs risks found in our study.

However, several limitations should also be noted. Firstly, we only measured the blood mtDNAcn in a single time-point at baseline, which may not reflect mtDNAcn variation over a lifetime and therefore might result in an attenuated estimation. However, Campa et al. measured two-time point mtDNAcn over 15-year period among 96 participants showed satisfactory stability with a high intraclass correlation (ICC=0.60) (Campa et al. 2018). In addition, we measured the levels of mtDNAcn in peripheral blood but not in the target GICs tissues, but another study identified a relatively high correlation between mtDNAcn derived from colorectal tissues and their corresponding peripheral blood (*r*=0.659, *P*=0.038) (Qu et al. 2011). Secondly, the sample size in our study was suggested sufficient for overall analysis, but was limited for stratification and interaction analysis as well as joint effect estimation, especially for GC and EC cases. Further studies with larger sample size are needed to validate the current findings. Thirdly, our study did not collect information of *H. pylori* infection status, so we could not assess the effects of *H. pylori* on the association of mtDNAcn with GC. Finally, the current investigations were conducted only in a Chinese population, which might restrict the generalizability of the current findings to other ethnic populations. More epidemiology researches with larger sample size and diverse human races are needed to confirm our findings and the underlying biological mechanisms warrant further investigations by *in vivo* and *vitro* studies.

## Conclusions

In summary, the current prospective case-cohort study indicates that both low and high mtDNAcn in peripheral blood are associated with 47% to 181% increased risks of CRC, GC, EC and total GICs in a middle-aged and older Chinese population. The robustness *U*-shaped association pattern provides valuable clues for better understanding the biological roles of mitochondrial underlying GICs development, and implies the potential of mtDNAcn as non-invasive biomarker for GICs risk prediction.

## Data Availability

Data sharing is not applicable to this paper as the datasets generated needed to be confidential.

## Acknowledgements

The authors want to thank all volunteers and medical assistants from the Dongfeng Motor Corporation in Shiyan, Hubei, China.

## Authors’ contributions

**Xin Guan**: Conceptualization, Methodology, Writing-Original Draft, Writing-Review & Editing. **Mengying Li**: Conceptualization, Methodology, Formal analysis. **Yansen Bai**: Formal analysis, Methodology, Investigation. **Yue Feng, Guyanan Li, Wei Wei, Ming Fu and Hang Li**: Investigation. **Chenming Wang, Jiali Jie, Hua Meng, Xiulong Wu, Qilin Deng, Fangqing Li**: Methodology. **Handong Yang, Xiaomin Zhang, and Meian He**: Resources. **Huan Guo\***: Project administration, Conceptualization, Writing-Original Draft, Writing-Review & Editing, Funding acquisition. All authors critically reviewed the manuscript and approved the final version of the manuscript.

## Availability of data and materials

Data sharing is not applicable to this paper as the datasets generated needed to be confidential.

## Ethics approval and consent to participate

The study protocol was ethically approved by the Ethics and Human Subject Committee of Tongji Medical College, Huazhong University of Science and Technology. Informed consent was obtained from all participants.

## Consent for publication

The authors have consented to publication after having read the final manuscript.

---

STROBE Statement—Checklist of items that should be included in reports of ***cohort studies***

|  | Item No | Recommendation | Reported on |
| --- | --- | --- | --- |
| **Title and abstract** | 1 | (a) Indicate the study's design with a commonly used term in the title or the abstract<br>(b) Provide in the abstract an informative and balanced summary of what was done and what was found | **Title and abstract** |
| **Introduction** |  |  |  |
| Background/rationale | 2 | Explain the scientific background and rationale for the investigation being reported | **Introduction** |
| Objectives | 3 | State specific objectives, including any prespecified hypotheses | **Introduction** |
| **Methods** |  |  |  |
| Study design | 4 | Present key elements of study design early in the paper | **Study population** |
| Setting | 5 | Describe the setting, locations, and relevant dates, including periods of recruitment, exposure, follow-up, and data collection | **Study population, Case-cohort study design and covariates assessment** |
| Participants | 6 | (a) Give the eligibility criteria, and the sources and methods of selection of participants. Describe methods of follow-up<br>(b) For matched studies, give matching criteria and number of exposed and unexposed | **Study population, Case-cohort study design and covariates assessment** |
| Variables | 7 | Clearly define all outcomes, exposures, predictors, potential confounders, and effect modifiers. Give diagnostic criteria, if applicable | **Case-cohort study design and covariates assessment** |
| Data sources/measurement | 8* | For each variable of interest, give sources of data and details of methods of assessment (measurement). Describe comparability of assessment methods if there is more than one group | **Materials and methods** |
| Bias | 9 | Describe any efforts to address potential sources of bias | **Materials and methods** |
| Study size | 10 | Explain how the study size was arrived at | **Materials and methods** |
| Quantitative variables | 11 | Explain how quantitative variables were handled in the analyses. If applicable, describe which groupings were chosen and why | **Case-cohort study design and covariates assessment, statistical analysis** |
| Statistical methods | 12 | (a) Describe all statistical methods, including those used to control for confounding<br>(b) Describe any methods used to examine subgroups and interactions<br>(c) Explain how missing data were addressed<br>(d) If applicable, explain how loss to follow-up was addressed<br>(e) Describe any sensitivity analyses | **Case-cohort study design and covariates assessment, statistical analysis** |
| **Results** |  |  |  |
| Participants | 13* | (a) Report numbers of individuals at each stage of study—eg numbers potentially eligible, examined for eligibility, confirmed eligible, included in the study, completing follow-up, and analysed<br>(b) Give reasons for non-participation at each stage<br>(c) Consider use of a flow diagram | **General characteristics of study participants** |
| Descriptive data | 14* | (a) Give characteristics of study participants (eg demographic, clinical, social) and information on exposures and potential confounders<br>(b) Indicate number of participants with missing data for each variable of interest<br>(c) Summarise follow-up time (eg, average and total amount) | **General characteristics of study participants, Table 1, figure 1, table S1, figure S1** |
| Outcome data | 15* | Report numbers of outcome events or summary measures over time | **Table 2, figure 2, figure 3, figure 4, table S2, table S3** |
| Main results | 16 | (a) Give unadjusted estimates and, if applicable, confounder-adjusted estimates and their precision (eg, 95% confidence interval). Make clear which confounders were adjusted for and why they were included<br>(b) Report category boundaries when continuous variables were categorized<br>(c) If relevant, consider translating estimates of relative risk into absolute risk for a meaningful time period | **Table 2, figure 2, figure 3, figure 4** |
| Other analyses | 17 | Report other analyses done—eg analyses of subgroups and interactions, and sensitivity analyses | **Table S2, table S3** |
| **Discussion** |  |  |  |
| Key results | 18 | Summarise key results with reference to study objectives | **Discussion** |
| Limitations | 19 | Discuss limitations of the study, taking into account sources of potential bias or imprecision. Discuss both direction and magnitude of any potential bias | **Discussion** |
| Interpretation | 20 | Give a cautious overall interpretation of results considering objectives, limitations, multiplicity of analyses, results from similar studies, and other relevant evidence | **Discussion** |
| Generalisability | 21 | Discuss the generalisability (external validity) of the study results | **Discussion** |
| **Other information** |  |  |  |
| Funding | 22 | Give the source of funding and the role of the funders for the present study and, if applicable, for the original study on which the present article is based | **Support statement** |

\*Give information separately for exposed and unexposed groups.
**Note:** An Explanation and Elaboration article discusses each checklist item and gives methodological background and published examples of transparent reporting. The STROBE checklist is best used in conjunction with this article (freely available on the Web sites of PLoS Medicine at http://www.plosmedicine.org/, Annals of Internal Medicine at http://www.annals.org/, and Epidemiology at http://www.epidem.com/). Information on the STROBE Initiative is available at http://www.strobe-statement.org.

