## Supplementary materials for "Associations of mitochondrial DNA copy number with incident risks of gastrointestinal cancers: a prospective case-cohort study"

| **Table S1.** Age and gender distributions for participants in the base cohort and sub-cohort. | | | | | | |
| --- | --- | --- | --- | --- | --- | --- |
| Age | Gender | Base cohort (n=21658) | |  | Sub-cohort (n=1173) | |
|  |  | n | % |  | n | % |
| <50 | Males | 154 | 0.71 |  | 10 | 0.85 |
|  | Females | 419 | 1.93 |  | 21 | 1.79 |
| 50-59 | Males | 703 | 3.25 |  | 34 | 2.90 |
|  | Females | 5233 | 24.16 |  | 287 | 24.47 |
| 60-69 | Males | 6309 | 29.13 |  | 330 | 28.13 |
|  | Females | 4471 | 20.64 |  | 232 | 19.78 |
| 70-79 | Males | 2292 | 10.58 |  | 133 | 11.34 |
|  | Females | 1750 | 8.08 |  | 103 | 8.78 |
| ≥80 | Males | 195 | 0.90 |  | 11 | 0.94 |
|  | Females | 132 | 0.61 |  | 12 | 1.02 |

| **Table S2.** The associations between baseline characteristics and blood mtDNAcn among the subcohort participants (n=1173). | | | | | | | | | | | |
| --- | --- | --- | --- | --- | --- | --- | --- | --- | --- | --- | --- |
| Characteristics | Subcohort (n=1173) | | |  | Males (n=518) | | |  | Females (n=655) | | |
|  | n | β (95% CI) | *P* ^a^ |  | n | β (95% CI) | *P* ^a^ |  | n | β (95% CI) | *P*^a^ |
| Age (years)^b^ | 1173 | 0.021 (-0.023, 0.065) | 0.341 |  | 518 | 0.017 (-0.062, 0.096) | 0.671 |  | 655 | 0.023 (-0.029, 0.075) | 0.377 |
| Gender |  |  |  |  |  |  |  |  |  |  |  |
| Males | 518 | -0.160 (-0.228, -0.092) | <0.001 |  |  | - |  |  |  | - |  |
| Education |  |  |  |  |  |  |  |  |  |  |  |
| Primary school or below | 331 | reference |  |  | 134 | reference |  |  | 198 | reference |  |
| Middle school | 443 | -0.054 (-0.135, 0.027) | 0.189 |  | 198 | 0.051 (-0.078, 0.180) | 0.441 |  | 244 | -0.139 (-0.245, -0.034) | 0.010 |
| High school or beyond | 399 | -0.100 (-0.183, -0.016) | 0.020 |  | 186 | -0.060 (-0.190, 0.071) | 0.369 |  | 213 | -0.132 (-0.242, -0.022) | 0.019 |
| BMI (kg/m^2^) ^b^ | 1173 | 0.015 (-0.085, 0.115) | 0.768 |  | 518 | -0.019 (-0.196, 0.158) | 0.834 |  | 655 | 0.033 (-0.086, 0.152) | 0.587 |
| <18.5 | 22 | -0.173 (-0.413, 0.066) | 0.156 |  | 9 | -0.208 (-0.598, 0.183) | 0.297 |  | 13 | -0.149 (-0.450, 0.153) | 0.333 |
| 18.5-24.9 | 660 | reference |  |  | 293 | reference |  |  | 367 | reference |  |
| ≥25 | 491 | -0.026 (-0.092, 0.040) | 0.445 |  | 216 | -0.056 (-0.160, 0.048) | 0.29 |  | 275 | -0.002 (-0.087, 0.083) | 0.971 |
| Smoking status |  |  |  |  |  |  |  |  |  |  |  |
| Never | 840 | reference |  |  | 204 | reference |  |  | 636 | reference |  |
| Former | 126 | -0.021 (-0.142, 0.1) | 0.407 |  | 121 | -0.033 (-0.165, 0.099) | 0.623 |  | 5 | 0.362 (-0.114, 0.838) | 0.136 |
| Current | 207 | 0.044 (-0.060, 0.147) | 0.731 |  | 193 | 0.063 (-0.055, 0.181) | 0.293 |  | 14 | -0.153 (-0.440, 0.134) | 0.296 |
| Alcohol drinking status |  |  |  |  |  |  |  |  |  |  |  |
| Never | 857 | reference |  |  | 241 | reference |  |  | 616 | reference |  |
| Former | 77 | 0.045 (-0.048, 0.138) | 0.343 |  | 69 | 0.027 (-0.131, 0.185) | 0.738 |  | 8 | 0.449 (0.073, 0.825) | 0.020 |
| Current | 239 | 0.07 (-0.069, 0.209) | 0.323 |  | 208 | 0.061 (-0.049, 0.171) | 0.279 |  | 31 | -0.059 (-0.255, 0.137) | 0.555 |
| Physical activity, yes | 1061 | -0.069 (-0.179, 0.04) | 0.216 |  | 466 | -0.033 (-0.202, 0.136) | 0.701 |  | 595 | -0.100 (-0.243, 0.044) | 0.174 |
| Family history of cancer, yes | 37 | 0.087 (-0.097, 0.271) | 0.354 |  | 11 | 0.010 (-0.341, 0.361) | 0.955 |  | 26 | 0.121 (-0.091, 0.333) | 0.264 |
| Peripheral blood cell counts |  |  |  |  |  |  |  |  |  |  |  |
| WBC, ×10^10^/L | 978 | 0.109 (-0.116, 0.333) | 0.342 |  | 437 | -0.009 (-0.373, 0.355) | 0.962 |  | 541 | 0.198 (-0.085, 0.480) | 0.170 |
| Platelets, ×10^10^/L | 980 | 0.001 (-0.005, 0.007) | 0.775 |  | 441 | 0.003 (-0.007, 0.013) | 0.582 |  | 539 | 0.001 (-0.007, 0.007) | 0.920 |
| **Abbreviations:** mtDNAcn, mitochondrial DNA copy number; BMI, body mass index; WBC, white blood cell; CI, confidence interval. | | | | | | | | | | | |
| **Notes:** ^a^ *P* values were derived from generalized linear regression models with adjustment for age and gender, while BMI, education levels, lifestyles, family history of cancer and peripheral blood cell counts were included as independent variables respectively. Only age was adjusted in gender stratification analysis. | | | | | | | | | | | |
| ^b^ In generalized linear regression model, age and BMI were transformed as every 10 years and every 10kg/m^2^, respectively. | | | | | | | | | | | |

| **Table S3.** Sensitivity analysis for the associations between mtDNAcn quartiles and incident GICs risks with adjustment for peripheral blood cell counts. | | | | |
| --- | --- | --- | --- | --- |
| Type of cancer | Quartiles of peripheral blood cells mtDNAcn | | | |
|  | Q1 | Q2 | Q3 | Q4 |
| No. of sub-cohort | 244 | 250 | 247 | 229 |
| ***CRC (n=231)*** |  |  |  |  |
| No of cases | 72 | 35 | 45 | 79 |
| HR (95%CI) | 2.12 (1.33, 3.37) | reference | 1.42 (0.86, 2.33) | 2.54 (1.62, 4.01) |
| ***GC (n=120)*** |  |  |  |  |
| No of cases | 27 | 21 | 24 | 48 |
| HR (95%CI) | 1.40 (0.76, 2.59) | reference | 1.29 (0.68, 2.44) | 2.63 (1.52, 4.55) |
| ***EC (n=64)*** |  |  |  |  |
| No of cases | 14 | 10 | 16 | 24 |
| HR (95%CI) | 1.58 (0.67, 3.75) | reference | 1.75 (0.75, 4.10) | 2.57 (1.17, 5.66) |
| ***Total GICs (n=415)*** |  |  |  |  |
| No of cases | 113 | 66 | 85 | 151 |
| HR (95%CI) | 1.81 (1.25, 2.62) | reference | 1.42 (0.96, 2.08) | 2.57 (1.80, 3.65) |
| **Abbreviations:** mtDNAcn, mitochondrial DNA copy number; HR, hazard ratio; CI, confidence interval; CRC, colorectal cancer; GC, gastric cancer; EC, esophageal cancer; GICs, gastrointestinal cancers. | | | | |
| **Notes:** Weighted Cox proportional hazards models were adjusted for age, BMI, gender, smoking (smokers/never smokers), alcohol drinking (current/non-current alcohol drinkers), physical activity (yes/no), education status (primary school or below, middle school, high school or beyond), family history of cancer, platelet counts and white blood cell counts. | | | | |

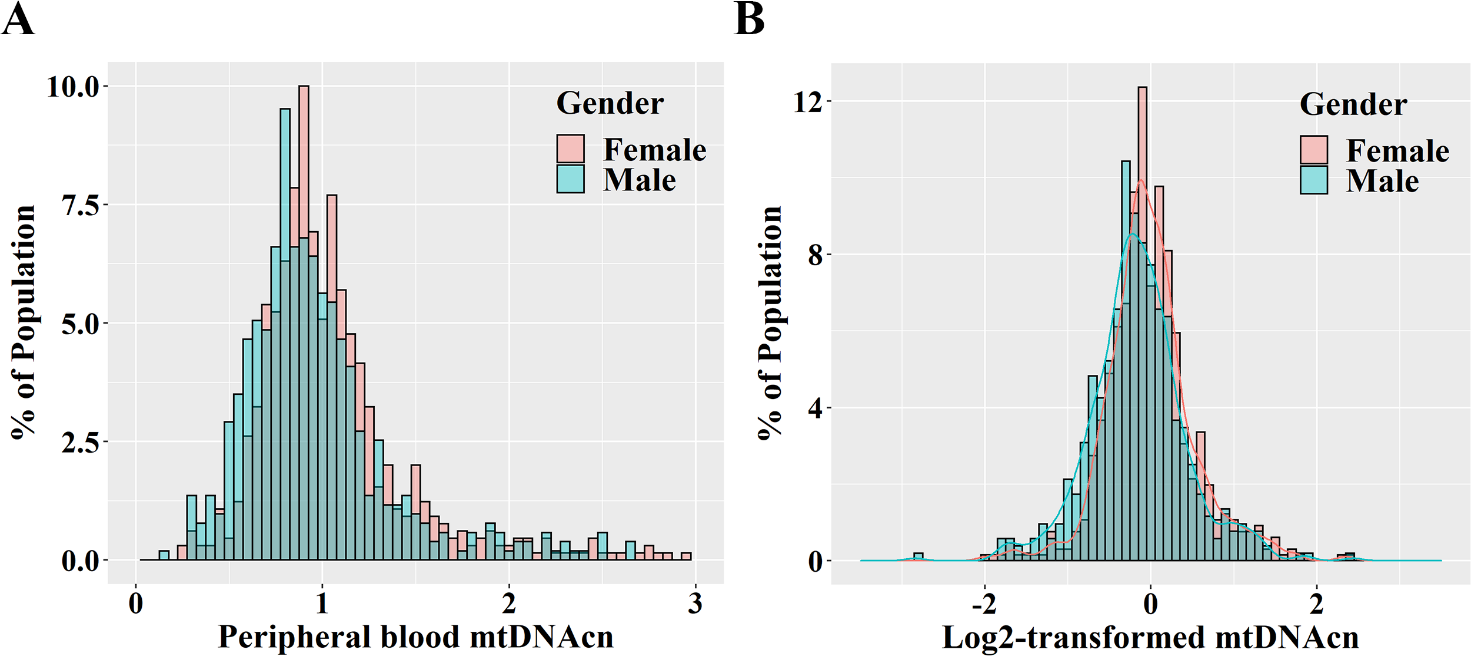

**Figure S1.** Distributions of peripheral blood mtDNAcn stratified by gender among subcohort participants (n=1173).

(A) Peripheral blood mtDNAcn (crude data);
(B) Log_2_-transformed peripheral blood mtDNAcn.

**Abbreviations:** mtDNAcn, mitochondrial DNA copy number.
